# Designing spatial adaptive surveillance for the emerging malaria vector *Anopheles stephensi* in Eastern and Horn of Africa

**DOI:** 10.64898/2026.03.05.26347695

**Authors:** Luigi Sedda, Eric Ochomo, Fitsum G. Tadesse, Bouh Adbi Khaireh, Assalif Demissew, Mulugeta Demisse, Dejene Getachew, Samatar K. Guelleh, Mohamed M. Ibrahim, Bernard Abongo, Vincent Moshi, Margaret Muchoki, Brian Polo, Janice Maige, Andrea M. Kipingu, Yeromin P. Mlacha, Onyango Sangoro, Monsuru Adeleke, Adedapo O. Adeogun, Babalola Ayodele, Fredros O. Okumu, Xiaoxi Pang, Heather M. Ferguson, Samson Kiware

## Abstract

The spread of *Anopheles stephensi* into the Horn of Africa represents one of the main challenges for malaria control, given the species’ ecological plasticity and resistance to multiple insecticides. In response to the World Health Organization’s 2022 vector alert, an adaptive, model-based spatial surveillance framework was developed and evaluated to improve detection, mapping accuracy, and operational responsiveness during invasion. Adaptive surveillance utilises initial observations to guide subsequent surveillance, linking the surveillance design to the underlying geographical characteristics of *Anopheles stephensi* distribution through observed data. This dynamic approach targets areas of high uncertainty and/or abundance, making the design responsive rather than predetermined. Focusing on Djibouti and selected regions of Ethiopia and Kenya, the adaptive surveillance was designed on previous in-country *Anopheles stephensi* surveillance data integrated with assembled open-source environmental, epidemiological, and demographic covariates. Key driver factors of the average monthly *Anopheles stephensi* catches varied geographically, although seasonality was universally important. Adaptive site allocation was optimised using a multicriteria target function which combines the trapping probability and uncertainty from previous surveys, with a simulation based on peaks-over-threshold (generalized Pareto) modelling of exceedances and Bayes factor–guided prioritisation. The selected adaptive surveillance design is the one that minimise the uncertainty in *Anopheles stephensi* trapping probability in hotspot areas. Optimal adaptive designs required between 50 to 59 sites per country, with uncertainty reductions in the probability of trapping projected up to 36% in Djibouti and more than 60% in Ethiopia and Kenya, with more than 60% site implementation halving uncertainty in Djibouti and Kenya and reducing it by up to 75% in Ethiopia. The proposed adaptive surveillance framework operationalises WHO guidance, accelerates hotspot identification, and inform targeted ecological studies and control interventions. It is extensible to other urban vectors (e.g., *Aedes aegypti*), enabling integrated, cross-border surveillance essential to contain *Anopheles stephensi* during ongoing invasion.

## Introduction

The recent expansion of the Asian malaria mosquito, *Anopheles stephensi*, beyond its traditional range marks a critical turning point in global malaria epidemiology. Its remarkable adaptability to both rural and urban environments threatens to undermine decades of progress in malaria control. This species’ ecological adaptability is compounded by its ability to oviposit in diverse larval habitats, including artificial water-storage containers and construction-related standing waters in urban areas, as well as streams, catch basins, wells, and domestic water-storage containers in rural settings [1, 2]. Control efforts are hindered by the tendency of *An. stephensi* to rest outside human dwellings and its resistance to all four major classes of insecticides commonly used for malaria control [3–5].

Historically, *An. stephensi* was confined to Southeast Asia, the Middle East, and the Arabian Peninsula [6]. Its first detection in Africa occurred in Djibouti in 2012 [6]. *Anopheles stephensi* was later detected in Ethiopia (2016), Sudan (2016), Somalia (2019), Somaliland (2019), South Sudan (2019), Nigeria (2020), Ghana (2022), Eritrea (2022), Kenya (2022), and the Republic of Niger (2024) [1, 3]. Some of these reports were delayed due to limited surveillance capacity in affected countries; especially for the emergence of new species [7].

The capacity of *An. stephensi* to thrive in urban areas and its competence in transmitting both *Plasmodium falciparum* and *Plasmodium vivax* have raised public health concerns in the African countries where it is emerging [8]. Malaria outbreaks in cities previously considered low-risk have been linked to this species, exemplified by the exponential rise in malaria cases in Djibouti following its initial detection [7]. Efforts to understand the ecological and epidemiological implications of *An. stephensi* remain constrained by the limited geographical coverage and scope of existing surveillance studies in Africa. Despite the ongoing expanded detection of this vector, there has been limited characterisation of its ecological adaptability and behavioural traits in African contexts. Invasion of new areas introduces uncertainties related to how this species is adapting to novel environments, host types, and local vector control measures. This paucity of region-specific data hampers accurate modelling of its distribution and impact on malaria transmission dynamics; limiting the development of contextually appropriate surveillance and intervention strategies [9].

In response to *An. stephensi*’s rapid geographic expansion in the part of the Horn of Africa, the World Health Organization (WHO) issued a ‘vector alert’ in 2022, urging intensified surveillance and implementation of specific control measures to halt the spread of this invasive malaria vector in Africa [7]. Most existing malaria vector surveillance designs in this continent rely on preferential surveillance guided by malaria risk or personal knowledge (e.g., [10–20]). These designs often lack long-term objectives, making them insufficiently aligned with the spatial and temporal dynamics of invasive malaria vectors [21, 22].

To address these gaps, this study developed an adaptive spatial surveillance design for *An. stephensi*, leveraging model-based geostatistics and extreme events theory [23] applied to existing *An. stephensi* surveys. This design accounts for the effect of inter-location distances on distribution map error [24], while maximising hotspot detection accuracy. Adaptive surveillance dynamically adjusts the surveillance strategy based on existing data, improving efficiency and effectiveness for achieving the surveillance objectives [25]. Stratification by malaria transmission risk and rural/urban land cover will enable estimation of *An. stephensi*’s relative contribution to urban and rural malaria, and identification of ecological and anthropogenic factors relevant for control [26].

Establishing and implementing an adaptive surveillance strategy is the essential first step to the WHO call, as it provides a flexible and efficient framework for rapidly detecting changes in vector distribution and guiding the design of detailed ecological and epidemiological studies during invasion events. In this study, we present an adaptive surveillance design for Djibouti, Ethiopia, and Kenya to maximise *An. stephensi* trapping probabilities and improve distribution mapping accuracy.

## Materials and Methods

### Study area

The study focused on parts of Eastern and Horn of Africa (in its broader definition), encompassing all of Djibouti, and selected regions of Ethiopia and Kenya (Supplementary Figure S1). For safety and political reasons, areas west of longitude 34.6°E (Ethiopia-Sudan border), and east of longitude 46.4°E (Ethiopia-Somalia/Somaliland border) were excluded, as well as regions south of latitude 2.40°S, corresponding to approximately 300km south of the latest detection of *An. stephensi* in Kenya, currently considered very low risk for occurrence of the species based on probability maps by Samake and colleagues [27]. Coordinates are expressed in the WGS84 geographic reference system.

#### Djibouti

The study area in Djibouti includes the city of Djibouti, its suburb of Balbala, five inland regions (Ali-Sabieh, Dikhil, Tadjourah, Obock, and Arta) and the localities of Wea, Douda, and Damjergo. In Djibouti malaria remains unstable with a resurgence between 2012 and 2020, following a steady decrease that started in 2008 taking the country to a pre-elimination level [28, 29]. Historically, *An. arabiensis* has been considered the primary malaria vector, along with *An. stephensi* in recent years. Malaria is mainly caused by *Plasmodium* falciparum, with a smaller proportion of *P. vivax*. Djibouti was the first African country of the three countries considered here to report *An. Stephensi* (September 2012, at the national animal export and quarantine station [30]).

#### Ethiopia

In Ethiopia, more than 70% of the population lives in malaria risk areas. The country has seen a resurgence of malaria in recent years with a 10-fold increase of malaria cases between 2019 and 2024. The principal malaria parasites are *P. falciparum* (around 60%) and *P. vivax* (around 40%) [8]. The main malaria vector is *An. arabiensis*, with *An. funestus*, *An. pharoensis* and *An. nili* considered secondary vectors [31]. In Ethiopia, *An. stephensi* was initially identified in Kebri Dehar in 2016, followed by detections in multiple urban settings, including Jigjiga (2018), Hawassa (2023) and Arba Minch (2024), one of the largest towns in southern Ethiopia [32, 33].

#### Kenya

The study encompassed 20 counties out of the total 47, including seven northern counties (Marsabit, Turkana, Mandera, Wajir, Isiolo, Samburu, Garissa, and Elgeyo Marakwet) where *An. stephensi* was detected previously [27]. Kenya exhibits heterogeneous malaria transmission across diverse ecological zones. One-third of Kenya’s population lives in high-risk areas. The main malaria vectors are *An. gambiae*, *An. arabiensis*, and *An. funestus sensu stricto*. Other species, such as *An. coustani* and *An. pharoensis*, also contribute to malaria transmission, with heterogenous importance across regions. Ten years after its detection in Djibouti and six years after Ethiopia, *An. stephensi* was reported in Kenya, initially in the northern region of the country, specifically in Marsabit and Turkana counties [34]. Subsequent targeted surveillance confirmed the widespread presence of *An. stephensi* in northern and north-central Kenya, including Isiolo, Elgeyo Marakwet, Mandera, Marsabit, Samburu, Turkana, and Wajir counties [27].

### Data

Prior to this adaptive surveillance plan, *Anopheles stephensi* trapping and surveillance strategies varied across countries, reflecting differences in surveillance objectives, logistical capacities, and epidemiological contexts.

In Djibouti, country-wide mosquito monitoring was conducted from December 2016 to March 2025, with collections carried out three days per week using Biogent BG-Sentinel traps baited with BG-Lure® attractant (BG Lure), and miniature Center for Disease Control and Prevention light traps (CDC LT). Key surveillance measures included mapping and sampling artificial water containers (plastic drums, tanks, ditches, manholes, wells) where larvae were found.

In Ethiopia, *An. stephensi* surveillance has evolved from *ad hoc* sampling to a more structured, longitudinal design in recent years. Surveillance, between November 2016 and January 2025, started in different localities once the invasive species was detected, especially in the eastern and northeastern parts of the country, including Somali, Dire Dawa and Afar regions. Later surveillance was intensified in the central part (Meki, Ziway and Metahara), and more recently in the southern (Hawassa and Arba Minch) and western parts of the country (Assosa). Enhanced surveillance targeted transportation corridors from Djibouti to the central and other regions of Ethiopia. Adult mosquito collection was conducted using animal/human baited tent traps, BG Sentinel traps (BG ST), black resting boxes, CDC Light Traps (CDC LT), clay pots, and mouth aspirators from animal sheds and later replaced with Prokopack aspirators. Larval collection focused on artificial water holding containers like cemented cisterns, barrels, plastic sheets, water drums around households, construction sites, and brick productions. A larger proportion of natural larval habitats were considered over time.

Finally, in Kenya surveillance was conducted between December 2022 and September 2024, targeting high-risk areas such as border regions, urban and peri-urban settings with suitable habitats, towns along major transportation routes, and counties reporting unusual malaria increases. Within selected counties, field teams prioritised locations with high livestock ownership due to the endo- and zoophilic behaviour of *An. stephensi* [5, 35], and with accessible water sources, including oases, dams, and construction sites. Both larval and adult mosquitoes were sampled, with adults trapped both indoors and outdoors. In most counties, sampling was conducted at three sub-counties, with two towns or villages surveyed per sub-county for eight days. Larval sampling targeted artificial and natural aquatic habitats such as water containers, irrigation ditches, and areas near animal shelters; with specimens preserved in ethanol. Adult mosquitoes were collected through quarterly cross-sectional surveys using BG lure, CDC LT, Human Landing Catch (HLC), Prokopack aspirators, and Ultraviolet light traps (UVLT).

The environmental, epidemiological, and demographic data used to model the average monthly *An. stephensi* catches were obtained from open sources (full description in Supplementary Table T1). Within these data, environmental variables known to have direct or indirect associations with *An. stephensi* presence [36], were obtained from remote sensing databases; epidemiological data (incidence of *P. falciparum* and *P. vivax*) from the Malaria Atlas Project; and human population data from NASA. Land cover was downscaled from 10m to 250m spatial resolution, assigning the modal land cover class to each grid node. Non-urban classes were reclassified as rural, creating a binary rural/urban variable. Malaria stratification followed the WHO Guidelines for Malaria [37]; with areas classified as having high transmission for *P. falciparum* and *P. vivax* if annual parasite incidence was of 450 or more cases per 1000 population; moderate for 250 to 450 cases, low transmission for 100 to 250 cases; very low when less than 100 cases per 1000 population were reported; and finally no transmission when no cases were reported in that year.

### Ethics statement

The use of secondary data, which contained anonymised mosquito household surveys, was approved by the Faculty of Health and Medicine Research Ethics Committee at Lancaster University (Reference Number FHM-2026-5742-ExRev-1). This study did not involve human subjects. All data pertained exclusively to households and contained no identifiable or individual-level human information.

### Statistical analyses

#### Selection of important explanatory variables

All variables listed in the Supplementary Table T1 were tested for collinearity prior to variable selection. All variables with a Pearson’s correlation coefficient above an absolute value of 0.6 were removed [38]. Two variables (Leaf Area Index and Enhanced Vegetation Index) were excluded, retaining those with higher significance, explained variance, and lowest AIC in univariate general linear models. For each country, variable selection involved testing all possible combinations of the remaining 14 explanatory variables excluding interactions (exhaustive search), within a spatial generalized linear model. The rural/urban variable and year were treated as a random effects and excluded from the selection process. The model with the lowest Watanabe–Akaike Information Criterion (WAIC) was considered to be the best-fitting model [39].

#### Statistical model

The response variable ***y*** was the *An. stephensi* catches with mean ***μ***.

We employed a spatial negative-binomial mixed model (SNBMM) with a log link function:

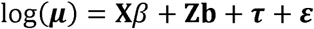

where the mean μ has covariance Δ; β is the vector of fixed effects coefficients for the design matrix of explanatory variables **X**; **b** is the vector of random effects coefficients for grouping variables (land cover and year) in the design matrix **Z**. **b** and the spatial dependent random errors, τ, and the spatially independent random errors, ε, have covariances:

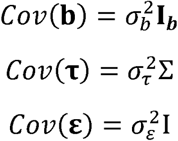

respectively; therefore the covariance of the observation under the mixed effects model is:

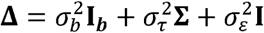

where 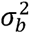 is the variance for the spatially independent random effects; 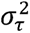 is the variance for the spatial dependent random errors, and 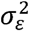 is the variance for the spatially independent random errors. ***I_b_*** is the identity matrix for the random effects; **Σ**[r] is the matrix determining the spatial dependence filled with distance-based weights, r, to adjust the spatially dependent variance; and **I** the identity matrix for the residual errors. In this study **Σ**[r] is a Matern correlation matrix [40], with correlation function for the matrix element *I*,*j* (representing two locations generically identified as *i* and *j*):

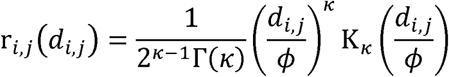

where K_κ_ is a modified Bessel function of the third kind and order κ (corresponding to a spatial field with *κ*-1 continuous derivatives), with *k* ∈ [0.2,5]. *d_i,j_*is the Euclidean where K_κ_ is a modified Bessel function of the third kind and order κ (corresponding to distance between locations *i* and *j*; and *ɸ* is the spatial range.

In addition to the explanatory factors described in Supplementary Table T1, the design matrix **X** includes the intercept; linear, quadratic and cubic seasonality effects (polynomial transformations of the time units); and for Kenya and Ethiopia, normalised larval presence probability γ*’* within a neighbourhood *D*:

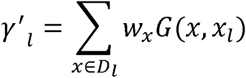

where *l* is a country grid node location, *w* the count of larva presence within a neighbourhood *D* from location *l*, and *G* is a Gaussian kernel [41]. A 1km neighbourhood has been used to consider mosquito maximum average dispersion.

Inference used a marginal approach with Laplace approximation [42, 43]. Predicted mean ***μ*** at location *i*, and model-inferred dispersion parameter, α, were used to estimate the trapping probability [44]:

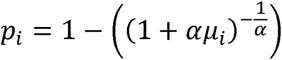

defined as the probability of trapping at least one *An. stephensi* during the observation period.

#### Model validation

Validation employed a single repetition 10-fold cross validation, averaging metrics across folds [45]. The employed validation metrics were: mean prediction error (bias), mean squared prediction error (MSPE), root mean squared prediction error (RMSPE), and the correlation between predicted and observed values. Low bias, MSPE and RMSPE, and a high correlation indicate good model performance. In the random selection of the locations for each fold, we have considered the potential bias caused by spatial autocorrelation (deflating the mean prediction error in presence of spatially clustered data), and therefore a buffering method to reduce spatial autocorrelation effects between training and test data [46, 47] was applied, with buffer size equal to spatial range, *ø*.

#### Adaptive surveillance

Adaptive sampling is a probabilistic preferential sampling where new units are added sequentially to an already existing sampling design according to a target function. The latter is based on the results from one or more models of the data obtained from the previous surveillance [22]. Therefore, the implementation of an adaptive surveillance design requires two components: an initial statistical model (described above) and a target function to inform where the surveillance sites need to be located. The target function is based on a multicriteria approach. The primary criteria identifies locations with the highest trapping probability for *An. stephensi* and the greatest uncertainty in prediction error. This improves hotspot delineation accuracy, i.e. identifying areas with the highest probability of collecting *An. Stephensi* mosquitoes. The secondary criteria ensures representativeness across rural and urban areas and malaria transmission risk strata.

The primary objective is maximised using peaks-over-threshold theory, where exceedances of mosquito trapping probabilities and uncertainties are assumed to follow a generalised Pareto distribution in space and time. To assess whether exceedance patterns for trapping probabilities and uncertainty are similar across space and time, we employed Bayesian factor analysis, comparing the plausibility of two competing models via marginal likelihoods. Bayes factor values between 0 and 2 indicate negligible differences. Therefore, minimizing the Bayes factor (no difference in the selected patterns of probability of trapping and uncertainty) becomes the objective in our adaptive surveillance design. This optimisation is achieved through simulation annealing applied to post-model inference results, iterating over batches of candidate locations of varying sizes. The method is fully described by Sedda and colleagues [23].

Once the spatial surveillance design is generated, locations are prioritised based on their capacity to improve hotspot delineation accuracy (largest accuracy improvement = highest priority), and to guarantee representativeness of urban and rural areas, and malaria transmission risk strata (secondary objective). To satisfy both criteria, some locations may fall outside high trapping probability zones.

## Results

The summary statistics of the field data used to produce the adaptive surveillance design are presented in Supplementary Table T2. In the three countries, adult *An. stephensi* average catches differed significantly (one-tailed Welch t-test, p-value ≈ 10^−4^), with Ethiopia reporting significantly larger catches (one-tailed Welch t-test, p-value ≈ 10^−7^). Model prediction errors were lower and with smaller confidence intervals for Djibouti and Kenya than Ethiopia (Table 1), likely due to the presence of isolated locations with very high catches in Ethiopia. The correlations between predicted and observed values were estimated between 0.6 and 0.8.

**Table 1.** Validation statistics for the SNBMM model. Metrics averaged across a 10-fold cross-validation. In brackets are shown the 95% confidence intervals.

| Validation statistic | Djibouti | Ethiopia | Kenya |
| --- | --- | --- | --- |
| Bias | 0.03 (-1.89,1.44) | 0.21 (-22.17,4.36) | 0.01 (-1.95,0.38) |
| MSE | 0.00 (0.00,3.00) | 0.57 (0.00,26.33) | 0.00 (0.00,2.31) |
| RMSE | 0.04 (0.01,1.73) | 0.75 (0.16,5.13) | 0.01 (0.00,1.52) |
| Correlation | 0.74 (0.49,0.80) | 0.79 (0.14,1.00) | 0.64 (0.24,1.00) |

Variable selection and their statistical significance within the SNBMM varied between countries. Water soil and vegetation content (MIR) were significantly associated with *An. stephensi* catches in Ethiopia, water vapour (NIR), vegetation and temperature in Kenya, with only elevation being important in Djibouti. Due to the limited number of explanatory factors for Djibouti, the variance explained by the predictors was the lowest among the three countries (Table 2). Notably, mosquito catches were associated with malaria parasite rates in the two countries where *An. stephensi* has been established the longest (Ethiopia and Djibouti); with larger odds in Ethiopia than in Djibouti (Table 2). Larval presence probability was a significant predictor of *An. stephensi* adult catches in Kenya but not in Ethiopia (the only two countries with larval collection).

**Table 2.**
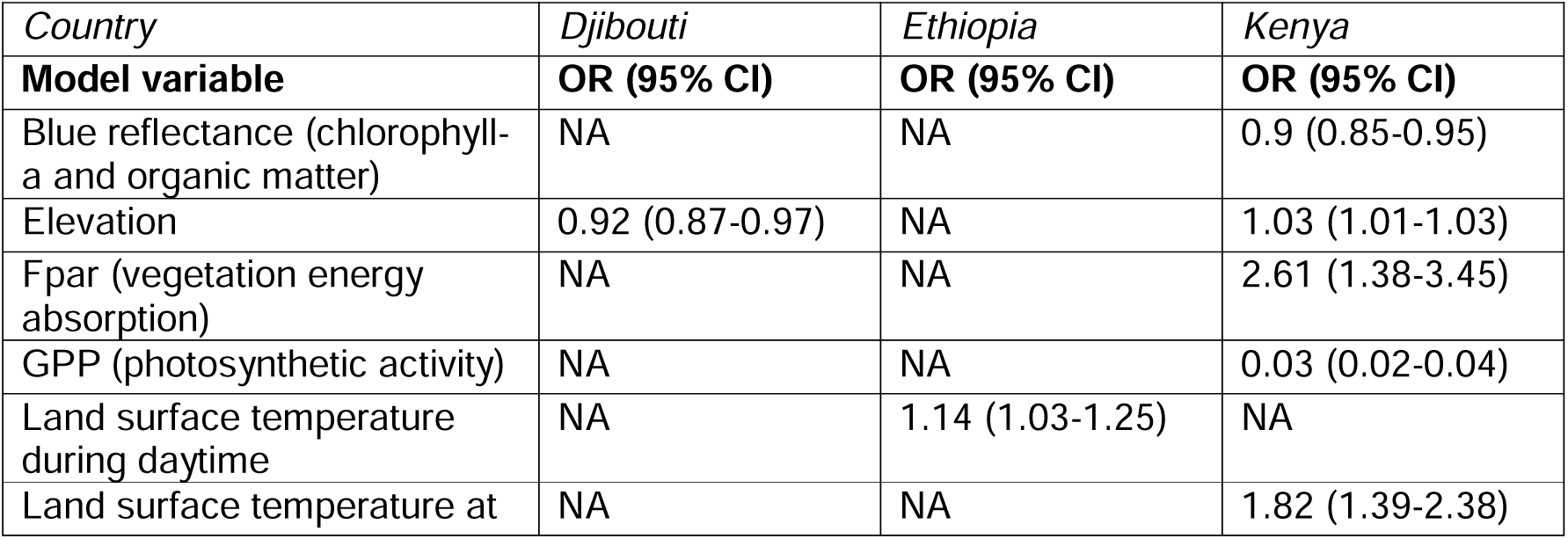
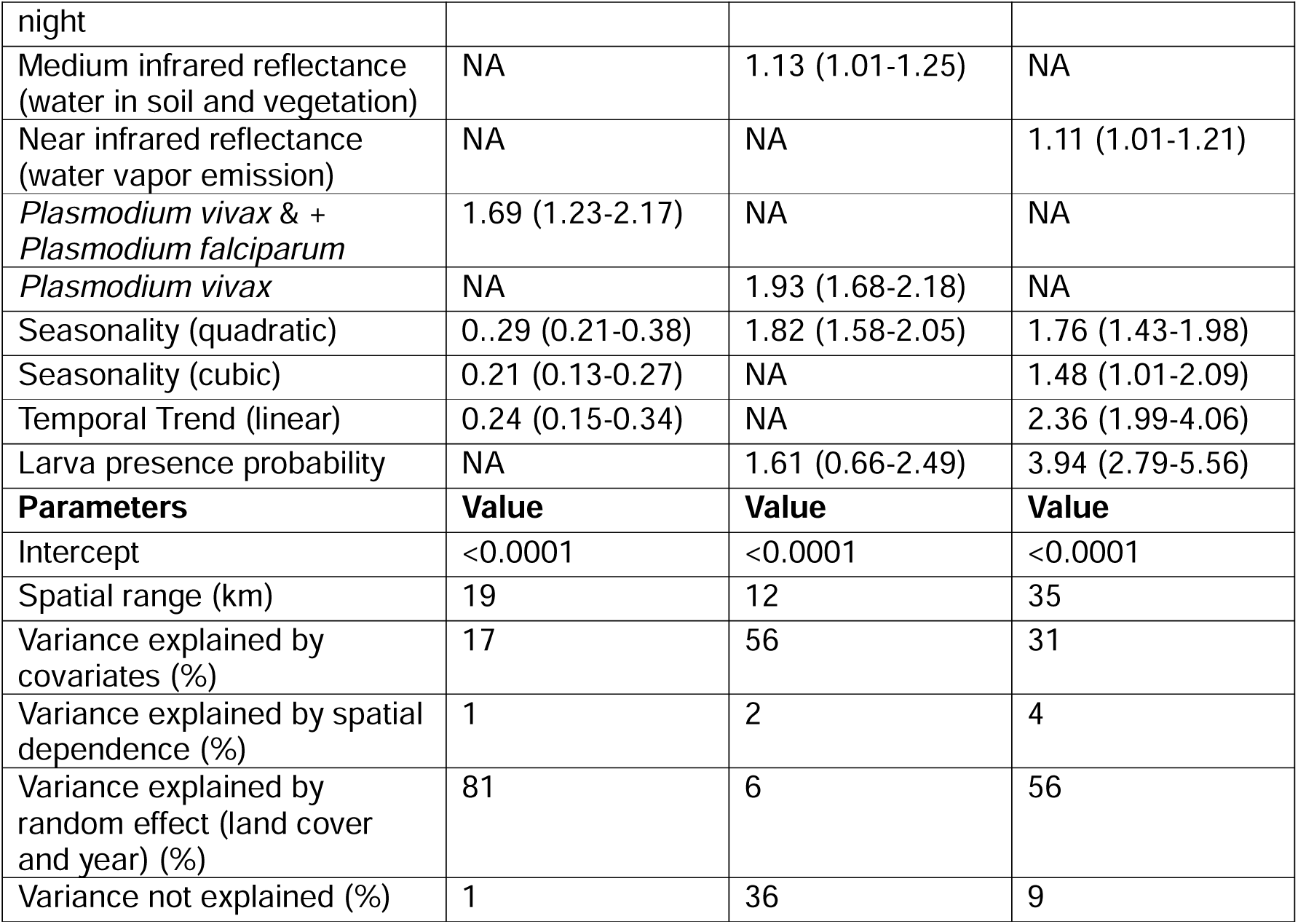
Odds ratios (OR) with relative 95% confidence intervals of explanatory variables in models of adult *An. stephensi* catches in the three African countries.

Quadratic seasonality (U or inverted U shape) was the only common significant variable across all country models, with a similar pattern of seasonality in Kenya and Ethiopia (U shape), but inverted in Djibouti. Other seasonal components were significant in Kenya and Djibouti, with Kenya showing increased catches over-time, opposite to Djibouti; and for the cubic seasonality one peak followed by one trough in Kenya, while one trough followed by one peak in Djibouti. From the analysis of the spatial autocorrelation, *An. stephensi* in Kenya appears to be characterised by a diffusion process due to the low catches but the largest spatial range of the spatially-dependent variance. However, in Djibouti and Ethiopia, the spatial range is below 20km, likely indicating mixed emergence and establishment phases.

The proposed adaptive designs include locations in both high and low trapping probability zones (Figure 1). Low-probability sites are often in proximity of high-probability areas or selected to reduce uncertainty from unsampled regions. To improve the distribution map, adaptive designs required between 50 to 59 locations per country, distributed across malaria risk transmission strata and urban/rural land covers (Supplementary Table T3). The allocation of these adaptive sites in Kenya required an increase to five urban sites to improve statistical representativeness.

**Figure 1.**
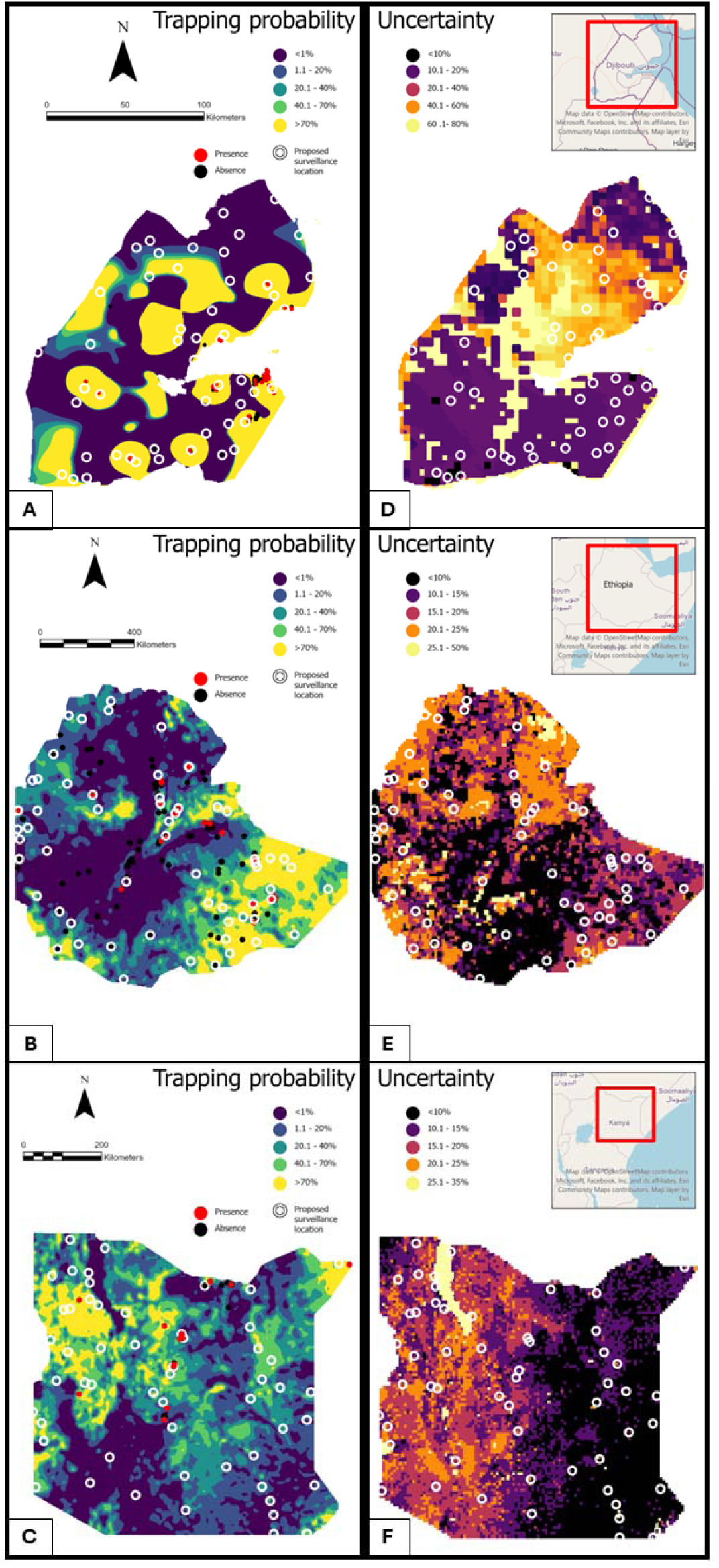
Trapping probability, proposed adaptive locations and historical *An. stephensi* presence or absence (from this study data) for Djibouti, Ethiopia and Kenya (A-C), along with uncertainty maps (D-F).

While the number of adaptive sites recommended by the model was similar for all three countries, their effect on the uncertainty is different. By deploying the adaptive surveillance in Djibouti a reduction of prediction uncertainty of up to 36% is expected, with reductions of above 60% for Ethiopia and Kenya (Table 3). The simulations showed that sampling more than 60% of the adaptive sites (around 30 to 35) can halve uncertainty in Djibouti and Kenya, and reduce it by up to 75% in Ethiopia.

**Table 3.** Effect of employing a proportion of the total number of adaptive surveillance sites (50 for Djibouti, 59 for Ethiopia and 56 for Kenya) on the uncertainty by country. Each country column shows the proportion of uncertainty reduction compared to the initial uncertainty (pre-adaptive) with relative 95% confidence intervals.

| % Adaptive locations | Djibouti | Ethiopia | Kenya |
| --- | --- | --- | --- |
| 10% | 6% (0%-20%) | 7% (4%-10%) | 7% (6%-8%) |
| 30% | 9% (0%-33%) | 13% (5%-21%) | 18% (17%-19%) |
| 60% | 15% (0%-50%) | 20% (5%-35%) | 31% (28%-34%) |
| 100% | 36% (6%-66%) | 62% (34%-90%) | 65% (35%-95%) |

## Discussions

The African expansion of *An. stephensi* poses a significant threat to malaria control progress in the Horn of Africa [48]. Given the complexity of its emergence, characterised by uncertain introduction routes and poorly understood bionomics, priority must be placed on strengthening surveillance systems. Enhanced surveillance will not only enable timely interventions but also accurate mapping, which is essential for assessing the mosquito’s impact on health and local economies [25]. Mosquito surveillance designs in Africa are typically purposive (e.g., targeting malaria high-incidence areas) or coverage-based [49]. Our framework offers strong methodological support for adaptive surveillance by providing flexible designs that can be optimised for additional objectives including accessibility and economic cost, and by quantifying accuracy gains from different design choices.

The validation results for the models informing adaptive surveillance, were satisfactory, with predictions aligning well with observed data. Our prior data show *An. stephensi* densities comparable to those reported in recent systematic reviews and meta-analysis [7]. Conclusions about environmental drivers of the *An. stephensi* catches in Africa cannot be elaborated accurately due to the species’ recent emergence and current environmental adaptation. However, our preliminary findings are broadly consistent with existing literature on the environmental drivers of *An. stephensi* (see below). The lack of significant medium-to-large scale environmental drivers in Djibouti may reflect the relatively stable climate conditions across the country, supporting year-round presence of *An. stephensi* [50, 51]. In fact, daytime surface temperature (mean 34.2 □C, standard deviation of 5.2 □C), and nighttime surface temperature (mean 24.01□C, standard deviation 3.5 □C), fall within the species’ suitable range [50–52]. Water (NIR) and temperature were significantly associated with trapping probability in Kenya. These results, along with not-significant human population density, were consistent with findings from Samake and colleagues’ *An. stephensi* suitability map [27].

In Ethiopia, despite previous studies reporting a weak link between *An. stephensi* seasonal predictability and precipitation [1, 32], we found significant association with seasonality and with surface water content (MIR) – although the latter does not only depend on precipitation. Notably, some adaptive sites were identified above 2000m elevation, a threshold for malaria-free zones established by the Ethiopian Ministry of

Health, but which are not completely malaria-free at the district level [3]. In addition, a low explained variance of rural/urban land cover (used as a random effect) suggests homogeneity in the abundance of *An. stephensi* across the country than previously thought. This was not confirmed by the models for Kenya and Djibouti [1].

Larval presence strongly predicted *An. stephensi* catches in Kenya, even when having a lower proportion of larvae-positive sites than Ethiopia, which suggested the presence of potential superproductive sites in Kenya [53]. Ability to identify such potential hotspots of larval productivity could greatly enhance the containment of *An. stephensi* through targeted Larval Source Management, particularly to stop further expansion where hotspots are identified near invasion fronts. The potential presence of superproductive sites is supported by *An. stephensi* clustered catches characterised by long range of spatial dependence within homogeneous environments [54]. In other words, the variation of adult catches is geographically dependent on the presence of larvae sites. In Kenya emergence is also characterised by a positive temporal linear trend in catches over the study period. However, in Djibouti the temporal linear trend is negative potentially due to a stronger effort in vector control. This confirms that the deployment of longitudinal studies in key *An. stephensi* hubs could be instrumental to assess its capacity to maintain stable populations.

Finally, the selected significant explanatory factors found in our study align with the findings from the first world *An. stephensi* suitability map by Sinka and colleagues [51], who identified temperature, surface water content and vegetation as the main drivers of *An. stephensi* suitability, along with human population density, although the latter not significant in our study. The lack of significance of human population density in the three countries compared to Asia is probably driven by the large number of *An. stephensi* surveillance sites in urban areas in Asia [55]. However, it is likely that environmental factors alone may not reliably predict seasonal variation of *An. stephensi* (see also [55] for rainfall), reinforcing the urgency of early detection and continuous monitoring. Major transportation routes, high cross-borders suitability manifested by the genetic connectivity between *An. stephensi* populations, and large populations at risk underscore the need for coordinated, cross-border surveillance and control efforts [27, 51].

Adaptive surveillance allows stakeholders to explore the impact of varying the number and spatial allocation of surveillance sites on the quality of trapping probability maps [56]. Beyond supporting public health managers, this framework can be integrated into citizen science initiatives, which are increasingly recognised as cost-effective, scalable, and sustainable solutions for mosquito surveillance and control, particularly in low-resource settings. Citizen science approaches can be enhanced by the use of spatial surveillance designs and open-source tools [57]. Once the current distribution of *An. stephensi* and its coregionalization with other autochthonous malaria vectors are understood, it will be possible to determine the species’ role in malaria transmission [58], identify areas at risk of colonisation [59], such as construction sites in rapidly urbanising regions [60], and develop integrated control. Future adaptive phases should also incorporate insecticide resistance in *An. stephensi* to strengthen control strategies [2].

This study has some limitations which influence the design of adaptive surveillance strategies. First, the use of different mosquito trapping and surveillance methods across sites introduces significant limitations to mosquito data analyses because each device has method-specific detectability and mosquito behavioural selectivity [61]. Even when the ‘trap type’ covariate is not statistically significant, residual bias may persist due to limited power, collinearity with environmental covariates, hierarchical shrinkage absorbing method effects into random components, measurement error in method classification, or effect heterogeneity across space and time that is masked in averaged models. Harmonisation of field protocols is therefore essential to mitigate potential method-induced biases, even when formal significance tests do not flag them, although no single best trapping method exists [62]. Second, sparse data collection across space and time limits the ability to detect emerging hotspots, increasing the risk of false negatives. Finally, the restricted set of considered explanatory factors may overlook key environmental or anthropogenic drivers, constraining the predictive power of surveillance models.

Addressing these gaps requires optimisation of the surveillance design within interdisciplinary collaborations among researchers, public health institutions, and local communities. This need is urgent because even if eradication of *An. stephensi* from the Horn of Africa is unlikely (as seen in the persistence of *Aedes albopictus* outside the native Southeast Asia), elimination from areas where it is still emerging remains possible, as demonstrated for *An. gambiae* in Brazil and Egypt [63]. Although our designs focus on *An. stephensi*, they can be adapted to other vectors for malaria or vector-borne diseases, as for example *Aedes aegypti*, an urban species sharing similar ecological niches and breeding preferences. Considering both species aligns with the Global Vector Control Response (GVCR) 2017–2030, which advocates integrated vector control and cross-country collaborations [48, 64].

## Acknowledgement

We are grateful to the Djibouti, Ethiopia, Nigeria, and Kenya staff of National Malaria Elimination Programme, the Ministry of Health and research institutions, and to all the community leaders, community health workers and volunteers of the three countries. We also thank the administrative and technical staff at the Ifakara Health Institute (IHI, Tanzania), recipients of the funding, for their support in the project management.

## Data availability statement

The entomological data used in this study are available in the Zenodo open research repository at the following link: https://doi.org/10.5281/zenodo.18642425

## Contributors

Funding acquisition: SK, EO, FT, BK, AA, HF, LS

Conceptualization: LS, EO, FT, BK, AA, AK, FO, HF, SK

Methodology: LS

Data collection, validation and curation: LS, EO, FT, AD, MD, DG, SG, MI, BA, JM, VM, MM, BP, YM, BA, XP, OS, MA

Analysis: LS

Supervision: LS, SK, EO, FT, BK

Writing first draft: LS

Writing - review & editing: All authors

All authors reviewed and edited the manuscript. All authors had full access to all the data in the study and had final responsibility for the decision to submit for publication.

## Patient and public involvement

Patients and/or the public were not involved in the design, or conduct, or reporting, or dissemination plans of this research.

## Patient consent for publication

Not applicable.

## Supplementary Information

**Supplementary Figure S1.**
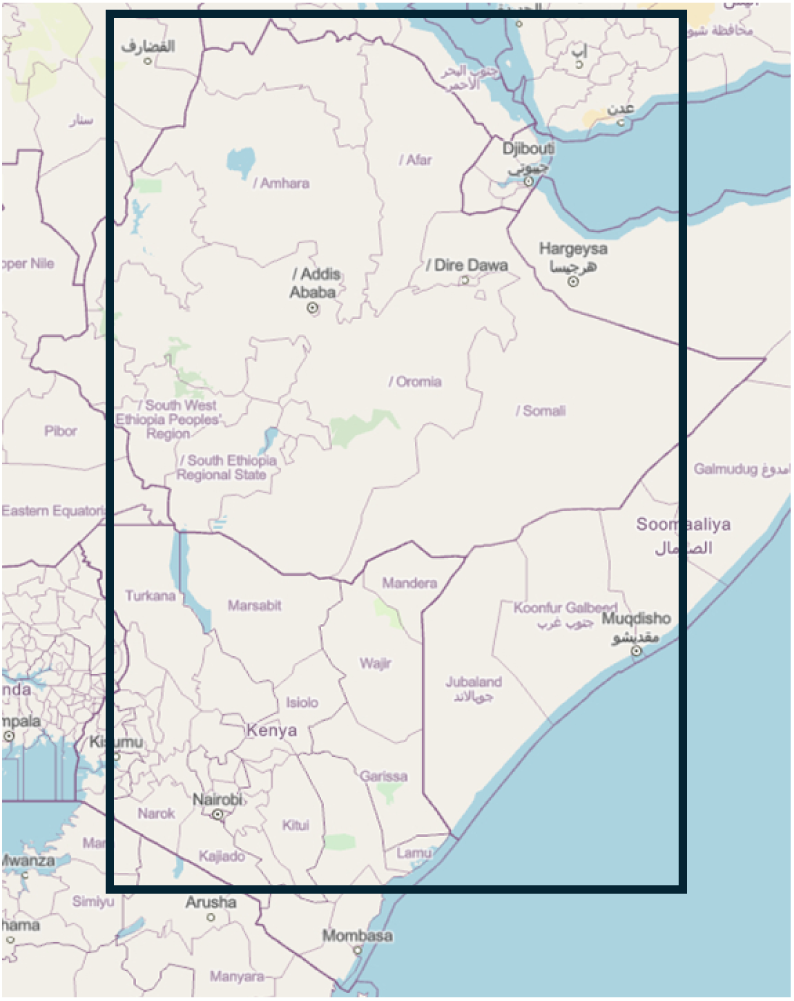
Study area. Map data © OpenStreetMap contributors, Microsoft, Facebook, Inc and its affiliates, Esri Community Maps contributors. May layer by Esri.

**Supplementary Table T1.** Data used for Anopheles stephensi modelling. All datasets (apart from human population) are relative to the same period of the mosquito collection.

| Variable | Definition | Spatial resolution (m) | Temporal resolution | Unit |
| --- | --- | --- | --- | --- |
| Blue<sup>1</sup> | Reflectance at 470 nm of the visible region of light spectrum– used for estimation of chlorophyll-a and organic matter. | 250 | 16 days | Dimensionless |
| EVI<sup>1</sup> | Enhanced Vegetation Index, a measure of vegetation growth, with improved sensitivity to high biomass regions compared to NDVI. | 250 | 16 days | Dimensionless |
| Fpar<sup>1</sup> | Fraction of Photosynthetically-Active radiation absorbed by vegetation as measure of vegetation energy absorption capacity. | 500 | 8 days | kilograms of Carbon per square meter |
| GPP<sup>1</sup> | Gross Primary Production – a measure of ecosystem photosynthetic activity. | 500 | 8 days | kilograms of Carbon per square meter |
| LAI<sup>1</sup> | Leaf Area Index to estimate vegetation canopy structure. | 500 | 8 days | Dimensionless |
| LST (day)<sup>1</sup> | Day Land surface temperature is a measure of near surface air temperature at daytime. | 1000 | 8 days | Kelvin |
| LST (night)<sup>1</sup> | Night Land surface temperature is a measure of near surface air temperature at nighttime. | 1000 | 8 days | Kelvin |
| MIR<sup>1</sup> | Middle Infrared Reflectance, 1629 nm, wavelengths prevalently absorbed by water, whether it is free water or water contained in plants. | 250 | 16 days | Dimensionless |
| NIR<sup>1</sup> | Near Infrared, 858 nm, measures water vapor. | 250 | 16 days | Dimensionless |
| NDVI<sup>1</sup> | Normalized Difference Vegetation Index (see EVI for differences). | 250 | 16 days | Dimensionless |
| PsnNet<sup>1</sup> | Net photosynthesis after removal of leaf and fine-root respiration. | 500 | 8 days | kilograms of Carbon per square meter |
| Red<sup>1</sup> | 648 nm of the visible region, often is used to measure darker and/or highly photosynthesised vegetation covers. | 250 | 16 days | Dimensionless |
| Elevation<sup>2</sup> | Elevation from a Digital Elevation Model named WorldDEM. | 90 | Yearly | meter |
| Land cover<sup>3</sup> | Annual Land Use Land Cover (9-class) V2<br>The 9 classes are: Water, Trees, Flooded vegetation, Crops, Built area, Bare ground, Snow/ice, Clouds, and Rangeland. | 10 | Yearly | Categorical |
| *Plasmodium falciparum*<sup>4</sup> | Annual incidence rate (number of newly diagnosed *Plasmodium falciparum* cases per 1,000 population, on a given year). | 5000 | Yearly | Case per person-time |
| *Plasmodium vivax*<sup>4</sup> | Annual incidence rate (number of newly diagnosed *Plasmodium vivax* cases per 1,000 population, on a given year). | 5000 | Yearly | Case per person-time |
| Human population<sup>5</sup> | Estimates of human population density based on counts consistent with national censuses and population registers with respect to relative spatial distribution, but adjusted to match the Revision of the United | 1000 | 2015 and 2020 | Number of persons per grid cell. |
|  | Nation's World<br>Population Prospects<br>(UN WPP) country<br>totals. |  |  |  |

Sources: <sup>1</sup> NASA: MODIS (https://planetarycomputer.microsoft.com/); <sup>2</sup> ESA: Copernicus (https://doi.org/10.5069/G9028PQB); <sup>3</sup> Impact Observatory, Microsoft, and Esri: ESA Sentinel-2 imagery (https://planetarycomputer.microsoft.com/dataset/io-lulc-annual-v02); <sup>4</sup> Malaria Atlas Project (https://data.malariaatlas.org/); <sup>5</sup> NASA (https://doi.org/10.7927/H4F47M65).

**Supplementary Table T2.** Sampling dates, sampling effort and summary statistics for the surveillance campaigns in Djibouti, Ethiopia and Kenya. M, male; F, female; T, total; SD standard deviation.

| Country | Djibouti | Ethiopia | Kenya |
| --- | --- | --- | --- |
| Time period (year: month) | 2016: 12<br>2022: 3,4,5<br>2023: 5<br>2025: 1,2,3,4 | 2016: 11<br>2018: 1,8<br>2019: 1,8,9<br>2020: 1<br>2021: 1,6,12<br>2022: 2,3,4,5,11<br>2024: 9,11<br>2025: 1 | 2022: 12<br>2023: 2,5,6,8,10<br>2024: 2,6,9 |
| Number of individual locations | 176 | 266 | 442 |
| Sampling effort (total number of trapping events) | 1805 | 310 | 465 |
| Number of collected *An. stephensi* | F: 301<br>M: 291 | T: 173 | T: 33 |
| Average number of collected mosquitoes | F: 0.16 (SD 0.57)<br>M: 0.17 (SD 0.57) | T: 2.33 (SD 6.65) | T: 0.07 (SD 0.59) |
| Collection method: sampling effort [average number of *An. stephensi*] | BG Lure: 860<br>[M=1.26, F=1.40]<br>CDC LT: 945<br>[M=0.16, F=0.17] | BG Lure: 3 [T=1]<br>CDC LT: 13 [T=1.54]<br>Black box: 3 [T=1]<br>Clay pot: 3 [T=0.33]<br>Animal/human baited: 18 [T=2.22]<br>Larval: 236<br>Aspirators/Prokopack: 16 [T=6.62]<br>Prokopack & CDC LT: 18 [T=0] | BG Lure: 28 [T=0.03]<br>CDC LT: 9 [T=1.33]<br>Larval: 22<br>HLC: 56 [T=0]<br>UVLT: 335 [0.03]<br>Prokopack: 15 [T=0.6] |

**Supplementary Table T3.** Stratification of the adaptive surveillance locations by malaria transmission risk and land cover. Classification definitions in main text.

| Stratum | Djibouti sample size achieved and [expected] | Ethiopia sample size achieved and [expected] | Kenya sample size achieved and [expected] |
| --- | --- | --- | --- |
| Malaria |  |  |  |
| *Moderate* | 8 [6] | 0 [0] | 0 [0] |
| *Low* | 20 [21] | 47 [46] | 45 [45] |
| *Very Low* | 22 [23] | 12 [13] | 11 [11] |
| Land cover |  |  |  |
| *Rural* | 42 [43] | 52 [52] | 51 [53] |
| *Urban* | 8 [7] | 7 [7] | 5* [3] |
| Total | 50 | 59 | 56 |

\* increased to improve statistical power of the stratum.

